# Use of causal DAG and regression analysis to understand and predict complicated osteoarticular infection in children

**DOI:** 10.1101/2025.06.26.25330329

**Authors:** Patrick H. Cahill, Aleisha Anderson, Daniel Yeoh, Matthew O’Brien, Timothy Robertson, Matthew Clifford, Cazz Finnucane, Andrew Martin, Katherine Stannage, Christopher Blyth, Julie Marsh, Asha C. Bowen, Tom Snelling, Charlie McLeod, Yue Wu

## Abstract

**Background:** While osteoarticular infections of the bones and joints in children usually resolve completely with adequate treatment, some children will develop complications. Predicting which children are at highest risk of complications might enable prevention through more aggressive treatment.

**Methods:** We used mutual information and Kullback-Leibler divergence methods to compare the characteristics of osteoarticular infections at the same tertiary institution in Australia in two time periods, 2002-2007 (N=295) and 2016-2018 (N=192). We used expert knowledge to develop a causal directed acyclic graph (DAG) that depicts the mechanistic pathways of osteoarticular infections and their progression to complications. Guided by the DAG, we developed three logistic regression (LR) models for predicting complications and evaluated their area under curve (AUC) to assess their performance for predicting complication.

**Results:** We observed a shift in the approach to diagnostic testing over the two time periods, with an increase in the number of blood cultures performed and a decrease in the rate of wound cultures. Fourteen of 43 test PCR tests (33%) for *K. kingae* recorded positive results. The established causal DAG clarified how the underlying, latent and dynamic biological processes become manifest as data. Utilising only data available at the initial point of care, the best LR model identified an optimal feature set that achieved an AUC of 0.85 for predicting complications.

**Conclusions:** Supported by domain expert knowledge and data, causal and statistical approaches were combined to offer valuable insights for predicting progression to complicated disease for children with osteoarticular infections.

**Key messages:**

- This study facilitates the understanding of change in epidemiology and clinical management of paediatric osteomyelitis and septic arthritis by examining a prospective cohort of 295 cases (2016-2018) in comparison to a similar study collected at an earlier date (2002-2007) from the same facility.
- We present a clarified understanding of the mechanistic pathways of osteoarticular infections and their progression to complications through the development of a causal directed acyclic graph (DAG) based on previously published DAG, recently collected data and domain expert knowledge.
- We demonstrate an approach to combine causal DAG, BN-based predictions of the causative pathogen, logistic regression modelling, and recursive feature elimination to achieve optimal performance in predicting complicated disease.

## BACKGROUND

Infections of the bone (osteomyelitis) and joints (septic arthritis) are collectively termed osteoarticular infection and occur in 4.3 per 100,000 Australian children per annum (1). The incidence among children from regional areas is estimated to be higher (2). While most children respond well to antimicrobial therapy, approximately 20% of osteoarticular infections require surgery (3) or extended hospitalisation (4,5,6). Some children develop chronic pain and permanent disability (5).

Osteoarticular infections are typically bacterial. Certain bacterial pathogen(s) like methicillin resistant *Staphylococcus aureus* (MRSA) have been implicated in complicated disease (7), however a pathogen is only identified in approximately half of cases (6). *Staphylococcus aureus* accounts for ∼60% of culture positive cases across all ages (5). *Kingella kingae* is now recognised as an important pathogen in pre-school-aged children (8). The epidemiology of osteoarticular infections has changed over time, with antimicrobial resistant strains becoming more prevalent (9). Early pathogen identification helps to support appropriate antibiotic selection in a timely manner (10,3,11,8,12) in order to avoid poor clinical outcomes.

The pathogen causing osteoarticular infection is traditionally identified by culturing blood, wounds, bone or joint tissue and/or joint fluid; more recently molecular methods have become available using PCR. Blood cultures have low sensitivity due to the low frequency of detectable bacteraemia in osteoarticular infection (11,13,14). Bone, tissue and fluid cultures are more sensitive but require invasive sampling procedures (15). Superficial wound samples yield a high frequency of false positive culture results owing to contamination of the biological specimen with normal skin flora (16,17). Molecular methods can be more useful for identifying difficult-to-culture (fastidious) bacterial pathogens but are not uniformly available across different healthcare settings (18,19). Attempts to identify a causative pathogen(s) can be challenging, time-consuming, and is often ultimately unsuccessful. Antimicrobial therapy is usually commenced empirically prior to knowing the outcome of diagnostic tests.

Causal Bayesian Networks (BN) depict the assumed causal mechanisms which underlie the clinical problem with a DAG, quantifying the strength of each causal relationship with conditional probabilities informed by expert knowledge and/or data-driven algorithms (14). BNs are a potentially valuable method for aggregating available clinical and laboratory patient data to provide an optimal estimate of the causative pathogen of infection (20,21,22,23,24,25) at the point of care. This could potentially improve empiric antibiotic selection and therefore patient outcomes. A simple BN can be used to explicitly model the mechanism underlying infection (26,27,28): namely, the relationships between demographic variables, a latent (unobserved) pathogen, various signs and symptoms, and test results. We have developed a BN model previously that reliably predicted the causative pathogen(s) for children with osteoarticular infections (11).

This study aims to inform clinical management of osteoarticular infections in children by predicting who’s more likely to develop complications based on data available to clinicians at the time of initial hospital presentations. By comparing data collected at one hospital in two time-periods (2002-2007) (6) and (2016-2018), we describe the changing epidemiology and clinical management of osteoarticular infections. Guided by domain knowledge and data, we developed a causal DAG to depict the mechanism of osteoarticular infection, its management and complications. Upon clarification of the disease mechanisms and data generation process, we implemented a method to detect a robust set of features for predicting progression to complicated disease utilising both recently collected data and causative pathogens predicted by our previously published BN (11) in a regression model.

## METHODS

In this section, we describe the collection and utilisation of data in understanding the epidemiology and clinical management of paediatric osteoarticular infections presenting to the tertiary paediatric referral hospital in Perth, Western Australia, over two time-periods. Data was collected between 2002-2007 (denoted hereafter as OM2002) (6) and 2016-2018 as part of the Western Australian Register of Septic Arthritis and Bone Infections prospective cohort study (WARSABI).

### OM2002 dataset

OM2002 comprised children aged 3 months to <16 years old hospitalised at Princess Margaret Hospital (PMH) with a clinical diagnosis of acute haematological osteomyelitis between 1st of September 2002 and 31st of August 2007. Further details regarding the epidemiologic features of this cohorts are described by Martin et al (6).

### WARSABI dataset

The WARSABI cohort comprised children 0 to <18 years old hospitalised with osteoarticular infection at PMH or Perth Children’s Hospital (PCH) between the 18th of July 2016 and the 17th of July 2018. Eligibility criteria included children with confirmed or presumed osteoarticular infection based on consistent symptoms and signs with or without supporting laboratory and radiological evidence. Prosthetic infections were included. Children who were found to have an alternative diagnosis (such as malignancy) were excluded.

A study nurse prospectively screened the emergency department admission log every weekday using the following terms: ‘osteomyelitis’, ‘arthritis’, ‘limping’, ‘pain’, ‘infection’, ‘effusion’, ‘ataxia’, and ‘bursitis’. Cases were also identified by notification of culture confirmed cases directly from laboratory or healthcare staff to a study investigator. Potentially eligible participants were provided with a hardcopy participant information sheet and written informed consent was obtained from guardians and participants ≥7 years old.

Clinical data were collected on standardized case report forms and entered into a secure electronic database (REDCap) hosted at PMH/PCH (29,30). Participants provided demographic, presentation, prior treatment and comorbidity details at study entry. Subsequently, clinical and treatment data including surgical procedures were obtained from the medical records, and microbiological data and radiological results were obtained from the hospital’s clinical information system. Children were followed up until 12 months to ascertain clinical and functional outcomes.

Cases were classified as community-acquired or healthcare-associated and as surgical site infections according to previously defined criteria (31). Infections were categorised according to the duration of symptoms before presentation: acute (*<*2 weeks), sub-acute (2 weeks to *<*3 months), or chronic (≥3 months).

### The Bayesian network model

The BN tested in this paper was trained using the data from OM2002, and its development has been detailed previously (11). This BN models the relationship between patient demographics, culture results and the sensitivity/specificity of those culture results with respect to the underlying latent causative pathogen of the infection. The structure of the model is shown in Figure 1.

**Figure 1:**
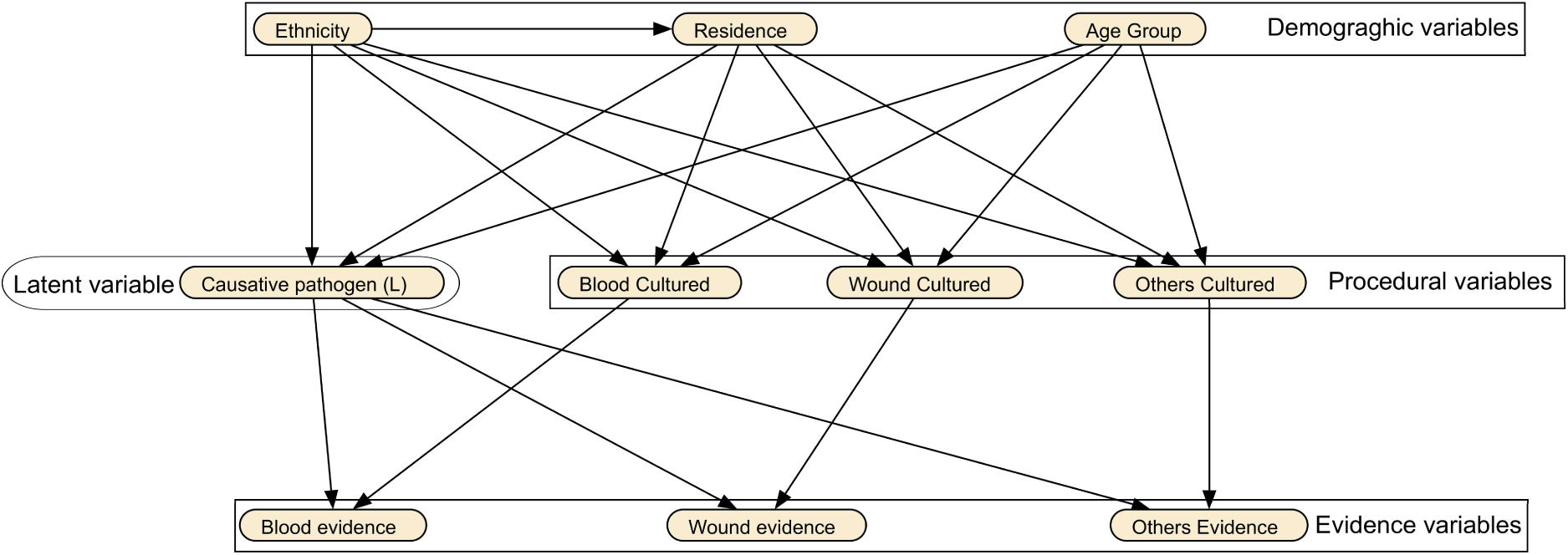
BN for osteoarticular infections.

Data variables collected from WARSABI participants were categorised into two broad categories: *BN variables* and *Other variables*, *i.e.*, variables referenced by the BN versus those which were not, respectively. For a given participant, the computed probability of each causative pathogen was conditioned on the observed values of demographic and laboratory BN variables for that participant. These probabilities were denoted *BN pathogen prediction*. Table 1 describes each BN variable and the prediction in detail, including any updates from (11), from which the model was adapted.

**Table 1:**
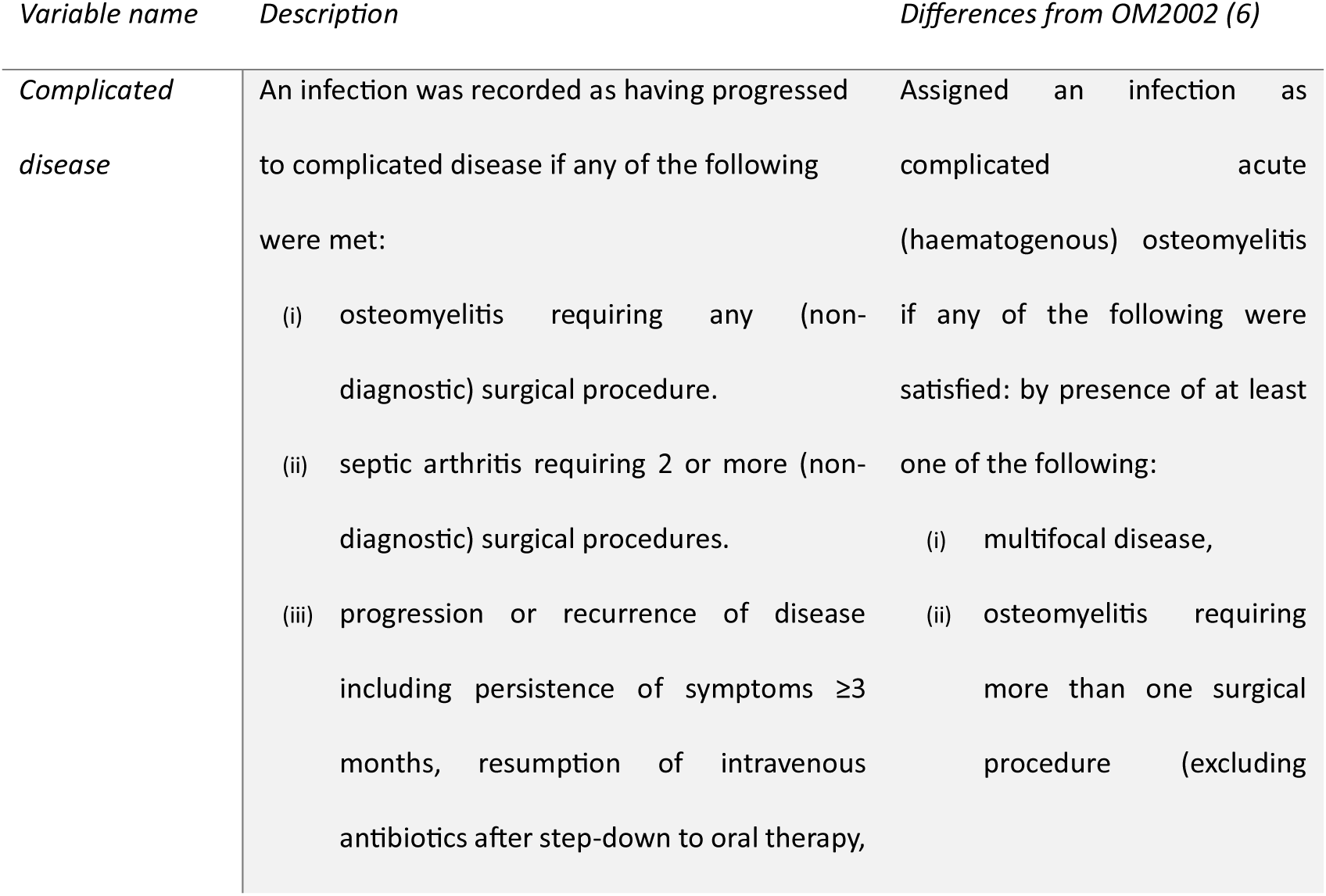
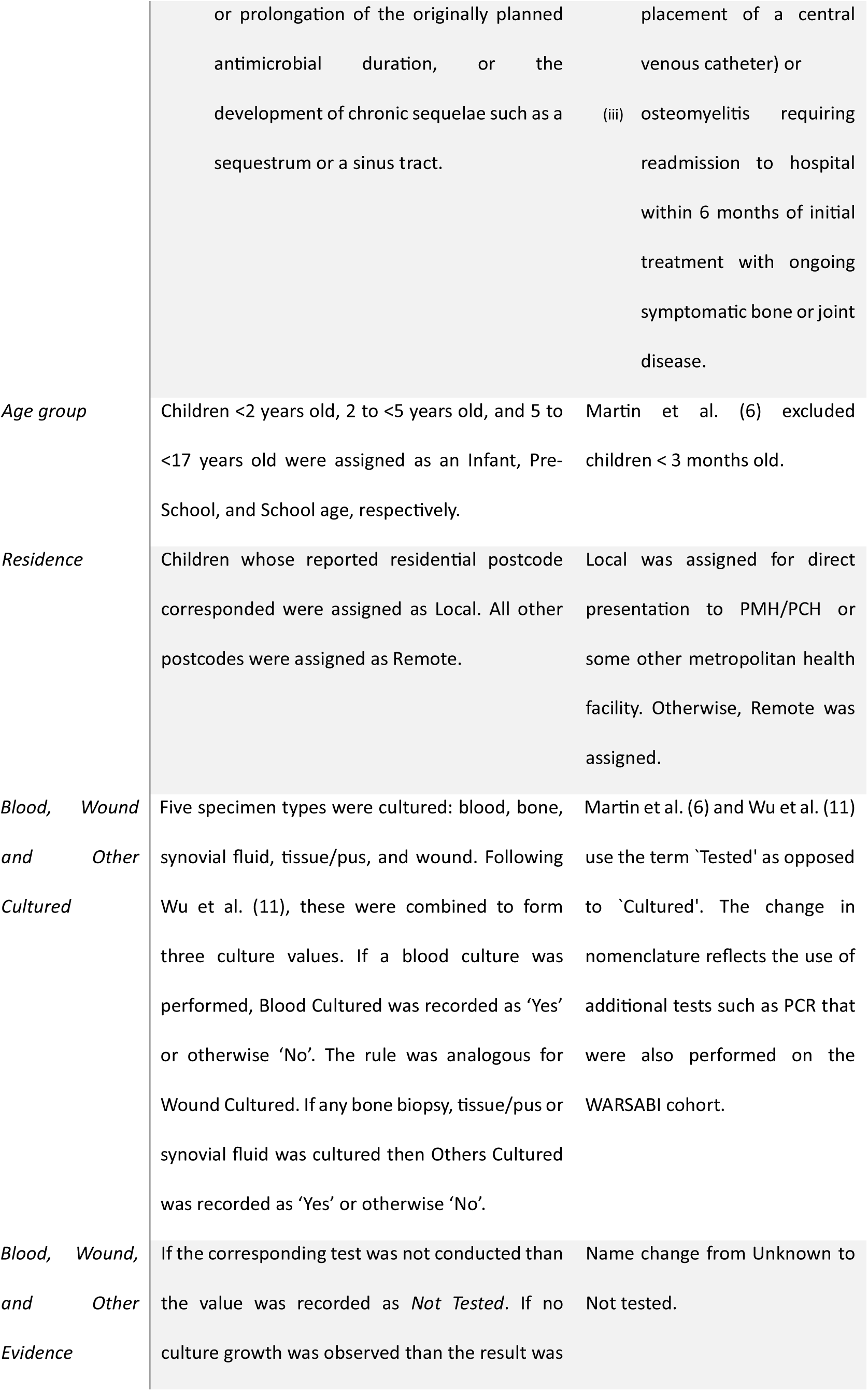
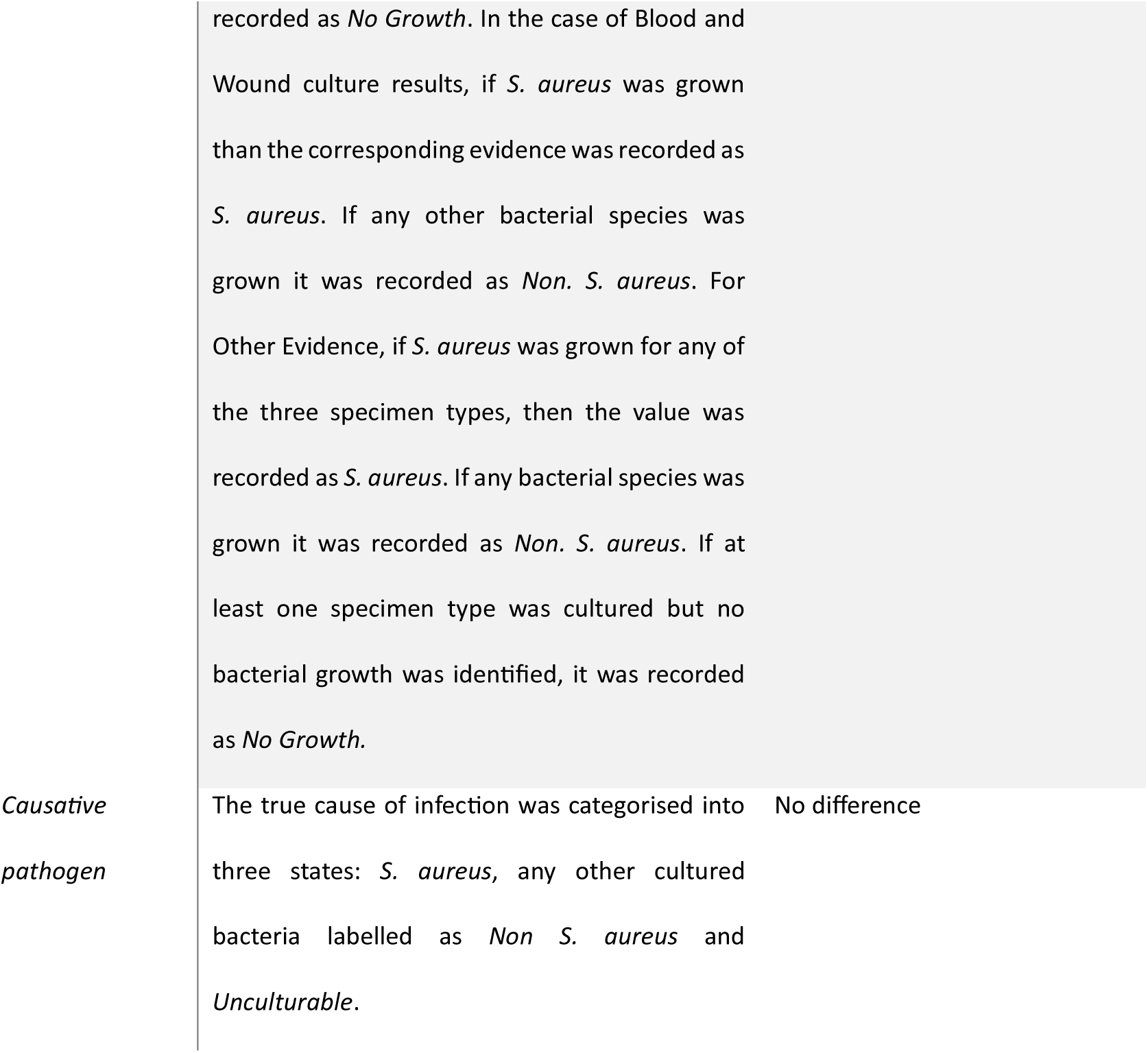
Complicated disease definition and BN variable definitions including any differences between WARSABI and OM2002.

| *Variable name* | *Description* | *Differences from OM2002 (6)* |
| --- | --- | --- |
| *Complicated disease* | An infection was recorded as having progressed to complicated disease if any of the following were met:<br>(i) osteomyelitis requiring any (non-diagnostic) surgical procedure.<br>(ii) septic arthritis requiring 2 or more (non-diagnostic) surgical procedures.<br>(iii) progression or recurrence of disease including persistence of symptoms $\geq 3$ months, resumption of intravenous antibiotics after step-down to oral therapy, | Assigned an infection as complicated acute (haematogenous) osteomyelitis if any of the following were satisfied: by presence of at least one of the following:<br>(i) multifocal disease,<br>(ii) osteomyelitis requiring more than one surgical procedure (excluding |
|  | or prolongation of the originally planned antimicrobial duration, or the development of chronic sequelae such as a sequestrum or a sinus tract. | placement of a central venous catheter) or (iii) osteomyelitis requiring readmission to hospital within 6 months of initial treatment with ongoing symptomatic bone or joint disease. |
| *Age group* | Children <2 years old, 2 to <5 years old, and 5 to <17 years old were assigned as an Infant, Pre-School, and School age, respectively. | Martin et al. (6) excluded children < 3 months old. |
| *Residence* | Children whose reported residential postcode corresponded were assigned as Local. All other postcodes were assigned as Remote. | Local was assigned for direct presentation to PMH/PCH or some other metropolitan health facility. Otherwise, Remote was assigned. |
| *Blood, Wound and Other Cultured* | Five specimen types were cultured: blood, bone, synovial fluid, tissue/pus, and wound. Following Wu et al. (11), these were combined to form three culture values. If a blood culture was performed, Blood Cultured was recorded as 'Yes' or otherwise 'No'. The rule was analogous for Wound Cultured. If any bone biopsy, tissue/pus or synovial fluid was cultured then Others Cultured was recorded as 'Yes' or otherwise 'No'. | Martin et al. (6) and Wu et al. (11) use the term 'Tested' as opposed to 'Cultured'. The change in nomenclature reflects the use of additional tests such as PCR that were also performed on the WARSABI cohort. |
| *Blood, Wound, and Other Evidence* | If the corresponding test was not conducted than the value was recorded as *Not Tested*. If no culture growth was observed than the result was | Name change from Unknown to Not tested. |

|  |  |
| --- | --- |
|  | recorded as *No Growth*. In the case of Blood and Wound culture results, if *S. aureus* was grown than the corresponding evidence was recorded as *S. aureus*. If any other bacterial species was grown it was recorded as *Non. S. aureus*. For Other Evidence, if *S. aureus* was grown for any of the three specimen types, then the value was recorded as *S. aureus*. If any bacterial species was grown it was recorded as *Non. S. aureus*. If at least one specimen type was cultured but no bacterial growth was identified, it was recorded as *No Growth*. |
| *Causative pathogen* | The true cause of infection was categorised into three states: *S. aureus*, any other cultured bacteria labelled as *Non S. aureus* and *Unculturable*. |

### Changes in disease profile

In the absence of a consensus definition for ‘complicated’ osteoarticular infection, we adopted a pragmatic definition defined in Table 1 that is similar to previous published definitions (6,32) and which captures clinically important outcomes.

With a robust network structure, small departures in the underlying epidemiology of osteoarticular infection from the training set should not significantly affect the performance of BNs (33), but large departures could do so especially if the BN conditional probability tables (CPTs) are not re-learned (23). Therefore, we compared the distribution of the baseline variables between OM2002 and WARSABI by investigating their Kullback-Leibler (KL) divergence and by assessing the changes in the mutual information between those variables in each cohort. We considered the mutual information matrix of the OM2002, WARSABI and synthetic data drawn from the BN distribution as well as a random distribution for comparison. The synthetic dataset was produced from the BN described in (11) using the Netica (https://norsys.com/) application to generate cases that matched the probability distribution. The random dataset was produced similarly with randomised CPTs. Each entry in the matrix is the mutual information between the variable in the column and the variable in the row. We present the ℓ^2^ norm distance between each mutual information matrix.

### Development of a causal DAG for osteoarticular infection

Adapting the structure of the BN presented in Figure 1, we developed a causal DAG to depict the mechanistic processes and clinical pathway for children with osteoarticular infection from initial hospital presentation to final follow-up. We identified 5 relevant time steps: a child with osteoarticular infection presents to the hospital at time *t*_−1_, is admitted to hospital at *t*_0_ and the results of their cultures and other tests become available at *t*_1._ The child is discharged from hospital at *t*_2_ and is followed until *t*_3_, say one year after discharge. Each variable was sorted into these time steps based on when data is likely to become available to the clinician. From this we constructed the model through expert consultation, including the following latent variables: "causative pathogen(s)" which corresponds to the true infecting organism (i.e. the aetiology), and "severity" of the inflammatory process at the different time steps. We also included "perceived pathogen" to represent the clinician’s evolving ‘working diagnosis’ of the causative pathogen. This informs their decisions regarding testing and treatment over time, possibly giving rise to a "change in antibiotic therapy". Although not captured in the datasets, it was considered important to understanding how beliefs and observed evidence inform management decisions. Supplementary Table S1 shows the definitions and corresponding WARSABI variables in detail. The DAG described in this paper were created using the GeNIe Modeler, https://www.bayesfusion.com/.

To investigate whether the *BN pathogen prediction* had value for predicting complications, we established three logistic regression models for predicting complications based on WARSABI data, each one only using data available at or before the time of admission, *t*_1_. The first model, denoted *L*_BN_, included only the *BN variables*, without access to the *BN pathogen prediction* The second model, denoted *L*_probs_, included only the *BN pathogen prediction* which represents the *BN variables* data processed by the BN, i.e. the estimated probabilities of each causative pathogen. Thirdly, we developed a model using all variables up to *t*_1_, i.e. both the *BN variables* and the *Other variables*. To avoid overfitting of this model, we employed recursive feature elimination (RFE) to identify a small subset of the most predictive variables (34,35). The mean AUC score across 10 iterations was calculated for 5-fold cross validated models trained on every available variable using an ‘lbgfs’ model with an *ℓ*_2_ penalty. We then recorded the mean score again with one variable removed and repeated this across every variable. We created a subset containing all the variables except the one that least reduced (or most improved) the performance. We repeated this process until no variables remained. We selected the feature set that had the optimal mean AUC-score across this elimination procedure and denoted this the optimal set bf_RFE_ and its corresponding logistic regression model as *L*_RFE_.

We report the receiver operating characteristic (ROC) curves and mean area under curve (AUC) results with respect to predicting complicated disease for the three models, as well as the mean model parameters, mean standard errors and corresponding feature odds ratios over each iteration. All categorical variables were dummy-encoded, and the data was scaled so each variable possessed a zero mean and unit variance. Reported coefficients and errors corresponded to the scaled values.

## RESULTS

### WARSABI epidemiology

Of the 192 children in the WARSABI cohort, 35% (N=67) were categorised as having complicated disease, of which 76% (N=51) were categorised as complicated because the patient required a single non-diagnostic surgical debridement.

### WARSABI and OM2002 data comparison

At least one risk factor was present in 93% (N=178) of cases in the WARSABI dataset but risk factors were not reported for OM2002 (6). In OM2002, comorbidities were recorded free-text rather than coded. **Error! Reference source not found.** shows a comparison of demographics, inflammatory markers, risk factors and comorbidities.

The proportion of children with a blood culture result was 66% (N=195) in OM2002 and 75% (N=144) in WARSABI; the proportion with a wound culture results was 23% (N=67) and 15% (N=29), respectively, and the proportion with culture of specimens other than blood was 24% (N=72) and 31% (N=59), respectively. The proportion of children with no culture results was 23% (N=69) and 16% (N=31), respectively.

In WARSABI, a PCR test for *K. kingae* was conducted in 22% (N=43) cases; of these, 32% (N=14) were positive. No PCR results were reported for OM2002 (6,11).

**Table 2:**
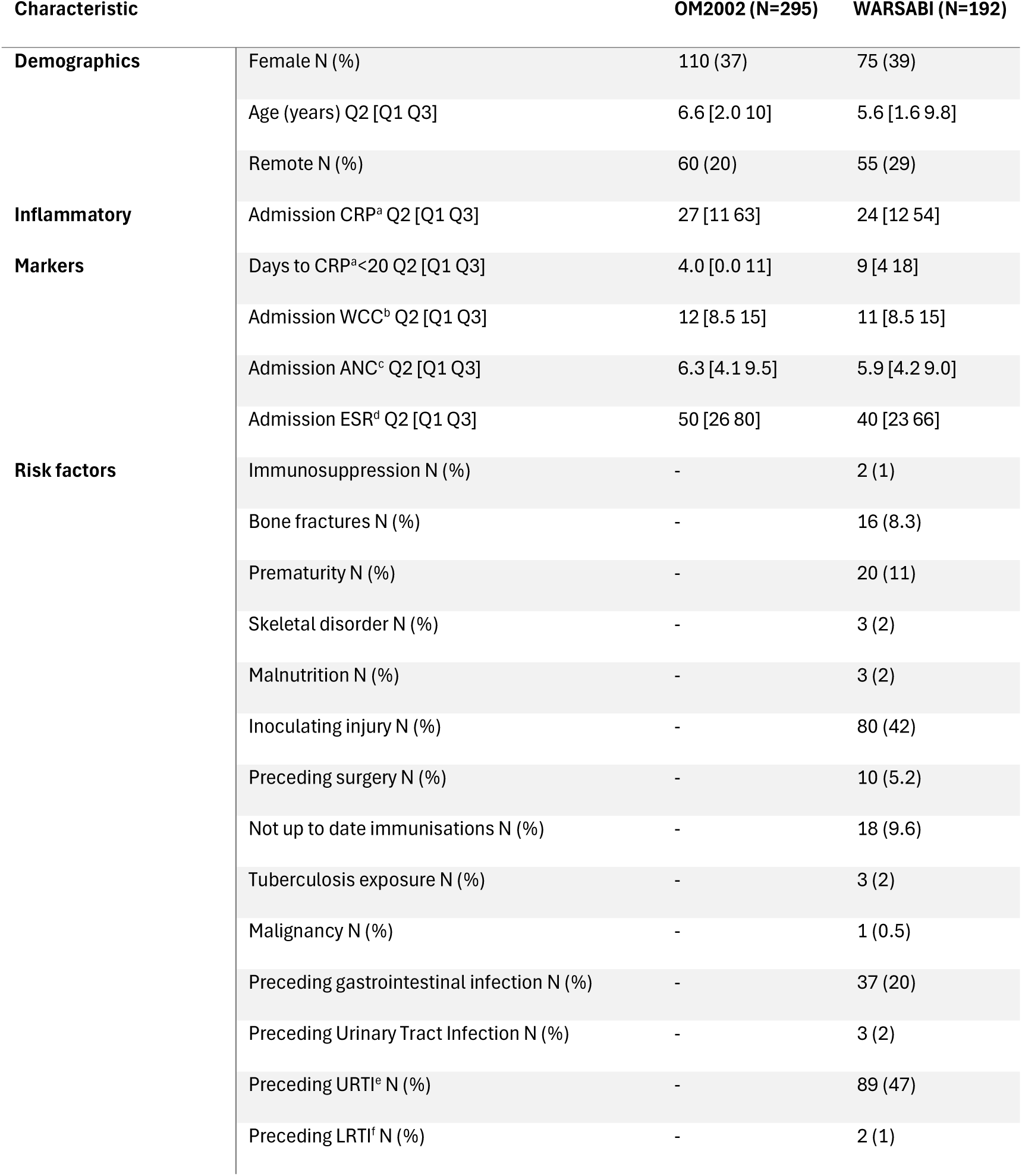
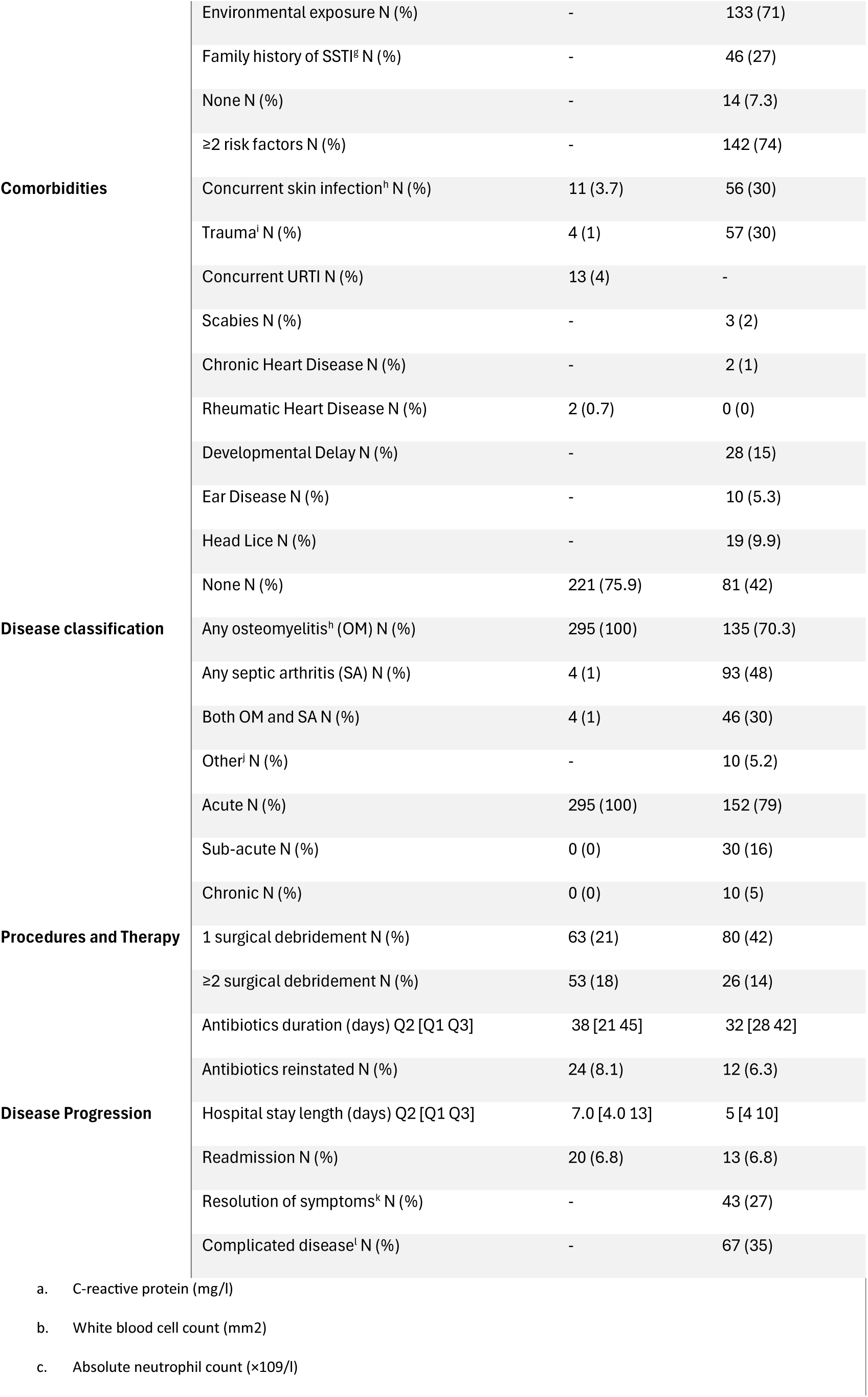
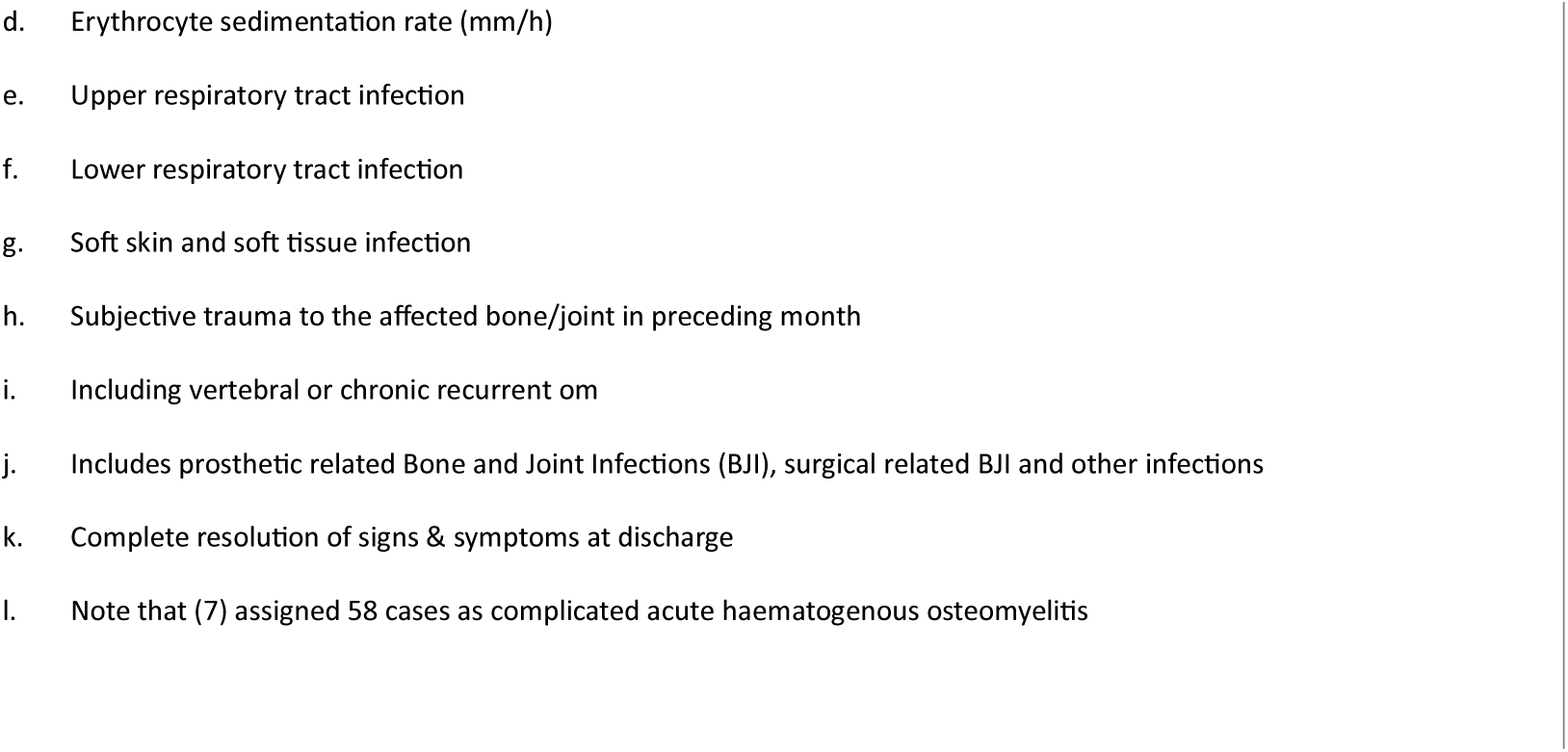
Comparison of the WARSABI data and the OM2002 data.

Figure 2 shows the mutual information and Kullback-Leibler divergence between the OM2002, WARSABI, synthetic and randomised datasets.

**Figure 2:**
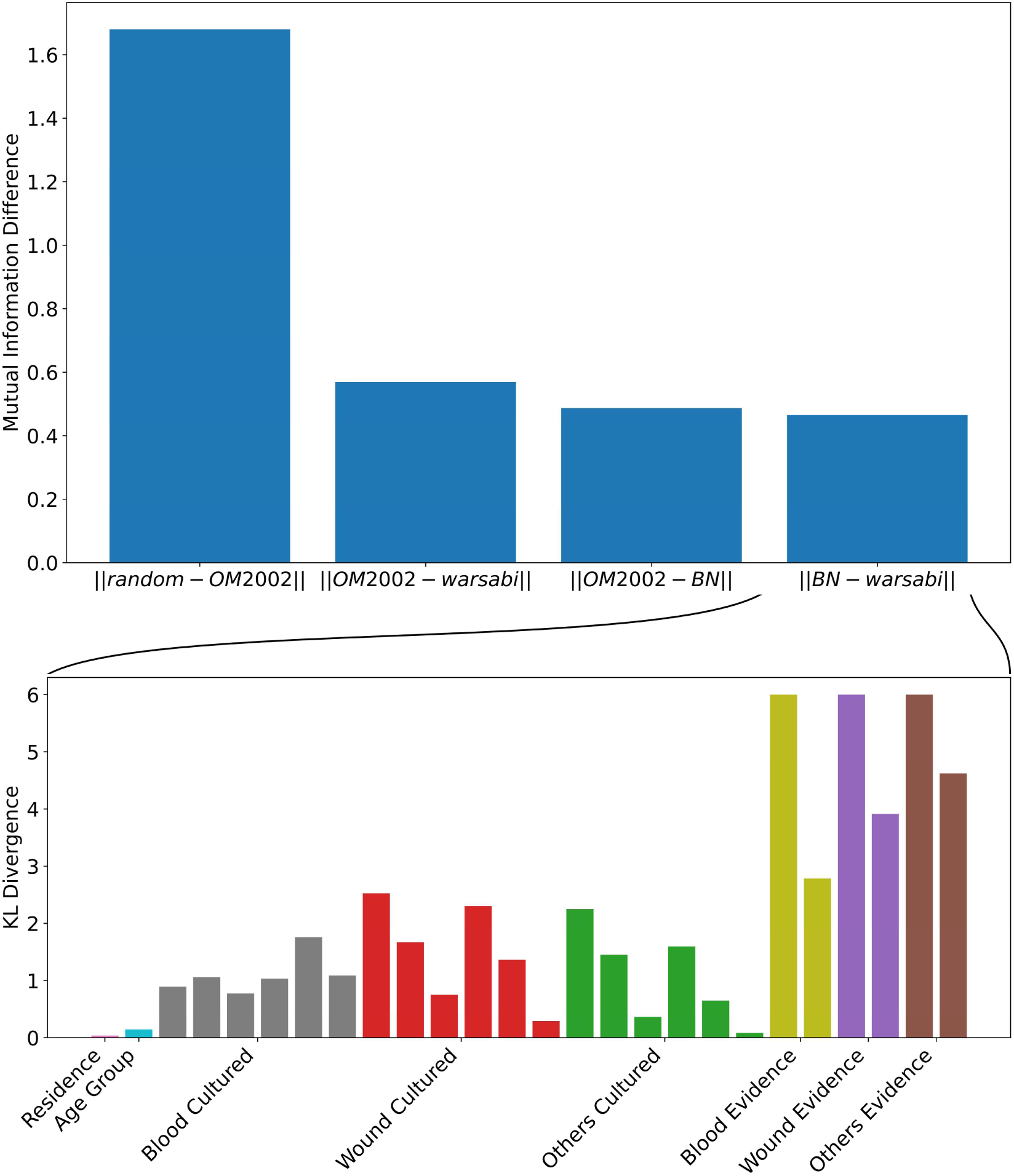
Comparison of the WARSABI data with the BN and the OM2002 dataset. **Top**: ℓ2 distance between MI matrices for BN, datasets and random BN. **Bottom**: Row KL divergence between WARSABI data and BN, using a log scale. Each column represents a different row of the CPT and is coloured according to which variable the CPT row corresponds.

### Causal DAG of osteoarticular infection

Figure 3 depicts the causal pathways underlying osteoarticular infection in children who present to hospital. The DAG consists of 96 variables in total, with 35 relating to background factors (blue), 11 relating to the mechanistic disease pathway (purple), 6 relating to management (pink), 33 relating to an overlap between disease and management (green), and 11 relating to disease outcomes (red). Complicated disease is defined by the status of the red outcome nodes.

**Figure 3:**
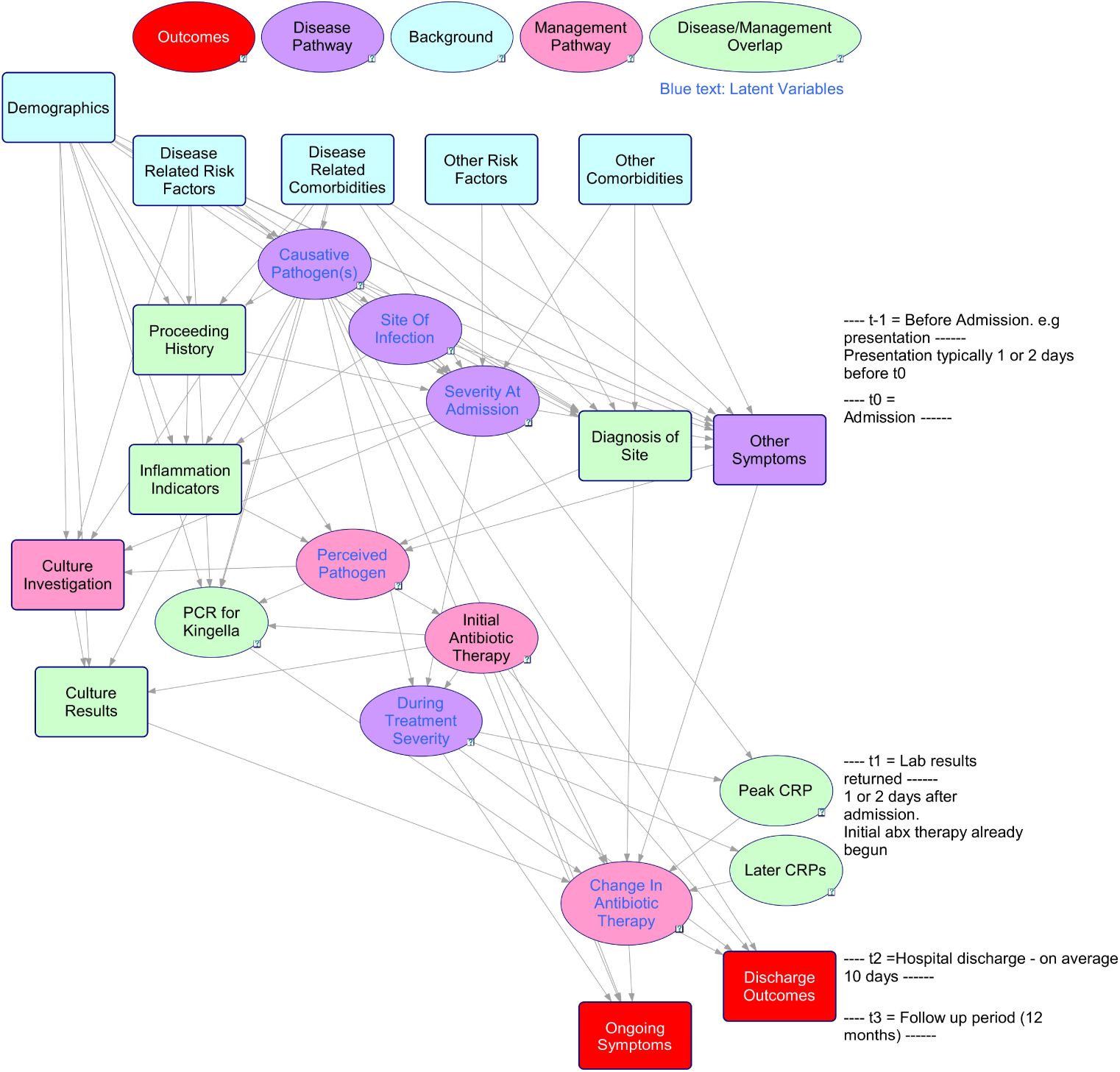
Causal DAG of osteoarticular infections, its management and complication.

### Data-driven predictors of complicated disease

We identified a set of 64 variables collected at (or before) *t*_1_ which are therefore likely to be available to the clinician at admission (Figure 3). See Table S1 for a full list and explanation of each variable included in this set. We performed recursive feature elimination as depicted in Figure 4 to produce an optimal feature set of variables for prediction of progression to complicated disease corresponding to the outcome sub-models occurring after *t*_1_ in Figure 3. We found that the optimal feature set, bf_RFE_, was: **Others evidence, admitting team, up-to-date immunisations, bone fractures, erythema of the overlying skin**, **joint immobility at admission and creatinine at admission**. Figure 4 shows the AUC score for each feature set by the feature set size. Similar results were observed when using an alternative (log-loss) scoring function to the AUC score (Figure S1). Table 3 presents the odds ratio and 95% confidence interval for each variable across the three models.

**Table 3:** Odds ratio (OR) with 95% confidence interval of *L*_RFE_, *L*_Probs_ and *L*_BN_ for each variable in each model. Variables are indicated in bold. For categorical variables, each value is indicated with italics.

| Variable Name | $L_{\text{RFE}}$ OR [95%] | $L_{\text{Probs}}$ OR [95%] | $L_{\text{BN}}$ OR [95%] |
| --- | --- | --- | --- |
| **Intercept** | 0.43 [0.24 0.70] | 0.46 [0.31 0.68] | 0.46 [0.28 0.76] |
| **Causative Pathogen** |  |  |  |
| **Probability** |  |  |  |
| *S. aureus* | - | 1.54 [1.30 1.81] | - |
| *Non S. aureus* | - | 1.12 [0.80 1.57] | - |
| *Unculturable* | - | 0.48 [0.34 0.69] | - |
| **Others Evidence** |  |  |  |
| *Not recorded* | 0.81 [0.42 1.80] | - | 0.84 [0.40 2.16] |
| *No Growth* | 0.55 [0.19 1.50] | - | 0.51 [0.18 1.43] |
| *Non S. aureus* | 1.03 [0.70 1.50] | - | 0.90 [0.60 1.37] |
| *Not Tested* | 0.69 [0.50 0.90] | - | 0.85 [0.73 0.99] |
| *S. aureus* | 2.07 [1.45 2.90] | - | 1.79 [1.23 2.6] |
| **Historical Bone Fracture** |  |  |  |
| *Yes* | 1.23 [0.89 1.70] | - | - |
| *No* | 0.82 [0.59 1.10] | - | - |
| **Up to date** |  |  |  |
| **Immunisations** |  |  |  |
| *Yes* | 0.81 [0.58 1.10] | - | - |
| *No* | 0.93 [0.65 1.30] | - | - |
| *Not recorded* | 1.75 [0.66 4.60] | - | - |
| **Joint Immobility** |  |  |  |
| *Yes* | 1.34 [0.94 1.90] | - |  |
| *No* | 1.36 [0.94 1.90] | - |  |
| *Not recorded* | 0.47 [0.25 0.90] | - |  |
| **Creatinine at admission** | 1.85 [1.14 3.00] | - | - |
| **Admitting Team** |  |  |  |
| *General paediatrics* | 0.76 [0.48 1.20] |  |  |
| *Orthopaedics* | 0.72 [0.49 1.00] |  |  |
| *Rheumatology* | 0.97 [0.51 2.20] |  |  |
| *Combined* | 1.08 [0.35 5.20] |  |  |
| *Plastics* | 1.97 [1.03 3.80] |  |  |
| *Other* | 1.21 [0.75 1.90] |  |  |
| **Age Group** |  |  |  |
| *Infant* | - | - | 0.90 [0.64 1.25] |
| *Pre-School* | - | - | 1.00 [0.70 1.42] |
| *School* | - | - | 1.12 [0.84 1.49] |
| **Blood Cultured** |  |  |  |
| *No* | - | - | 1.02 [0.89 1.17] |
| *Yes* | - | - | 0.98 [0.85 1.13] |
| **Blood Evidence** |  |  |  |
| *No Growth* | - | - | 0.92 [0.70 1.23] |
| *Non S. aureus* | - | - | 1.06 [0.50 2.88] |
| *Not Tested* | - | - | 1.02 [0.89 1.17] |
| *S. aureus* | - | - | 1.07 [0.76 1.52] |
| **Others Cultured** |  |  |  |
| *No* | - | - | 0.85 [0.73 0.99] |
| *Yes* | - | - | 1.18 [1.01 1.38] |
| **Residence** |  |  |  |
| *Local* | - | - | 0.91 [0.72 1.14] |
| *Not recorded* | - | - | 1.13 [0.65 2.33] |
| *Remote* | - | - | 1.08 [0.86 1.37] |
| **Wound Cultured** |  |  |  |
| *No* | - | - | 0.84 [0.74 0.96] |
| *Yes* | - | - | 1.19 [1.04 1.36] |
| **Wound Evidence** |  |  |  |
| *No Growth* | - | - | 1.11 [0.40 4.25] |
| *Non S. aureus* | - | - | 0.92 [0.64 1.34] |
| *Not Tested* | - | - | 0.84 [0.74 0.96] |
| *S. aureus* | - | - | 1.29 [0.93 1.80] |

**Figure 4:**
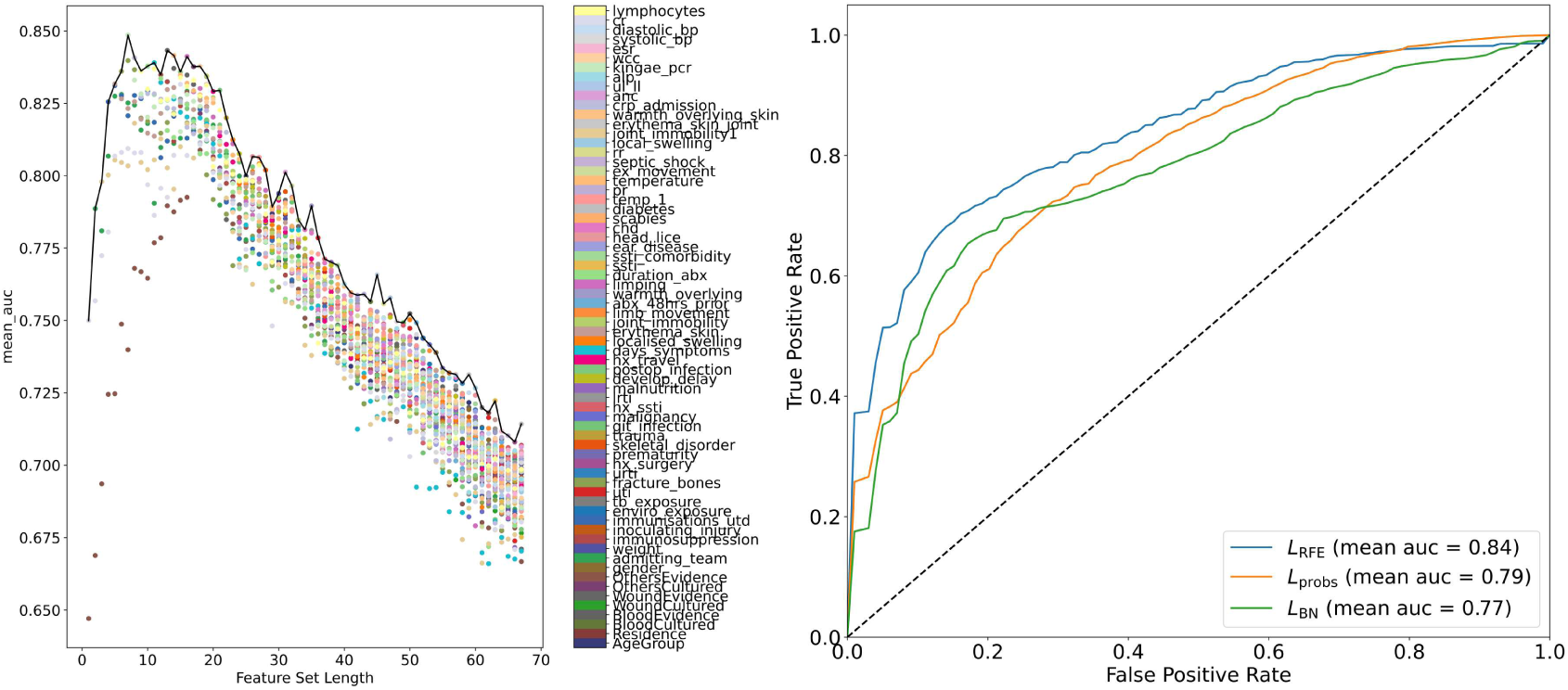
Recursive feature elimination results on WARSABI dataset. **Left:** Mean AUC over 10 iterations of 5-folds feature reduction leads to the following score against the size of the remaining feature set. Each scatter point gives the performance of the model after a specific variable was removed, while the black line shows the best performing feature set for each feature set length. The scatter points that lie on the line reflect the variable that was chosen for removal by the recursive feature elimination algorithm. **Right:** ROC curves comparing the performance of the model trained on the optimal feature set, L_RFE_; performance of the model trained on the BN variables, L_BN_; and the performance of the model trained on the causative pathogen probabilities, P_probs_.

## CONCLUSIONS

We presented differences in the characteristics of children admitted with osteoarticular infections to the same paediatric hospital in Australia over two time periods, approximately one decade apart. We have developed a causal DAG modelling the clinical pathway of an osteoarticular infection, which enabled the implementation of a new model for investigating the performance of Bayesian networks, using them to predict progression to complicated disease. We also implemented a method to detect a robust set of features for predicting progression to complicated disease.

The dynamic nature of this bacterial infection requires an understanding of the potential shifts in its epidemiology or clinical features over time. The demographic characteristics of patients in the WARSABI cohorts were similar to OM2002 and, although the proportion of cases from remote areas increased. Figure 2 (Figure 2) indicate that the differences between the BN and both the OM2002 dataset and the WARSABI dataset were less than the differences between the datasets themselves. This suggests that our BN, developed using expert-solicited opinion in combination with the OM2002 cohort data, effectively captured the relationships between the variables of interest, aiding the prediction of children at risk of disease complication at the time of admission.

Our causal DAG (Figure 3) proved useful in several ways. First, it helped clarify our definition of complicated disease (Table 1). For example, earlier draft definitions of included the presence of bacteraemia, however analysis of the DAG illustrates how this could only be realised if blood cultures were taken. Inclusion of blood culture result could overestimate the predictive value of our model. Secondly, it also helped to establish the set of variables used in the recursive feature elimination process by clarifying which variables (evidence) would be available to clinicians soon at the time of admission and therefore usable in a clinical context.

We found that incorporating the *BN pathogen prediction* into a logistic regression model for predicting complicated disease improved the model beyond use of the *BN variables* alone. This provides support to the validity of our causal DAG (Figure 3) which assumes a key role for the mostly latent (unobserved) causative pathogen in driving patient outcomes, and suggests the information encoded in the BN remains informative even after later observations become available and despite small departures from the training set relationships which may have occurred.

We implemented recursive feature elimination to create an optimal feature set of variables for predicting progression to complicated disease. We found that the optimal set improved upon a regression model comprising the *BN pathogen prediction* alone with an AUC=0.84 under 5-fold cross-validation compared to an AUC=0.79. Moreover, we suggest that the identified variables are not only predictive, they may be causally important. Immunosuppression, bone fracture, postoperative infection, preceding URTI and preceding antibiotics lie in the disease-related risk factors node of the causal DAG. Others evidence lies in the culture results node. Concurrent SSTI lies the disease-related comorbidities node. Joint immobility lies in preceding history. Respiratory rate lies in inflammation markers. Each node is a parent or a child of the latent severity at admission and the causative pathogen nodes. Both these unobserved nodes are hypothesised to drive patient outcomes and therefore, progression (or not) to complicated disease. It is plausible that some of the variables in the optimal set have predictive value because they provide evidence on these latent factors. While immunisation lies on the causal pathway, failure to document the immunisation status predicted complications rather than under-immunisation *per se*; the mechanisms which might explain the relationship between completeness of documentation and the risk of complications remain should be explored.

A limitation of the work presented here is that while we have provided evidence that the BN encodes relevant information, we have not quantified how much the epidemiology and management of osteoarticular infection has changed. The strength of this work is it provides interpretable measurements of the performance of a BN for modelling new data. Moreover, we have developed an effective method for producing an optimal feature set for the prediction of future outcomes. It has the advantage that it can be applied for any classification task that requires one to predict an outcome that occurs after admission, i.e after *t*_1_in the causal DAG in Figure 3.

In future work we will include treating the variables identified by the RFE process as candidates for inclusion in developing model-based clinical decision support tools for managing osteoarticular infections, for example, extending the structure of the simple BN as well as update the existing BN parameters using the WARSABI data.

## Ethics

Ethics approval for the data collection in this study was obtained from the Child and Adolescent Health Service Human Research Ethics Committee (2016032EP). Ethics approval for the development of relevant causal and statistical models was obtained from the Sydney Children’s Hospitals Network Human Research Ethics Committee (2020/ETH02510).

## Data availability

The data underlying this article cannot be shared publicly due to the privacy of individuals that participated in the study.

## Supplementary data

Supplementary data may be available upon request, subject to ethics approval.

## Author contributions

PC, YW, TS and CM contributed to the design of this project. CM, JM, AM, CB, AB, AM, KS and TS contributed to the design of the WARSABI study and CM led this study. TR, MO, AA, CF, MC, KS, CB, AB, TS and CM were involved in the implementation of the WARSABI study. PC and YW performed the analyses for this project. PC drafted the initial manuscript. All authors contributed and have approved the final version of the manuscript.

## Use of artificial intelligence tools

No AI tools were used to produce this article.

