## Supplemental Tables S1-3 and Figure S1 for "Use of causal DAG and regression analysis to understand and predict complicated osteoarticular infection in children"

### S1 Dictionary of terms

Table S1 shows explanations for terms (with abbreviated terms where applicable) used in the main text and their corresponding terms in the WARSABI data dictionary.

*Table S1: Dictionary of variable terms used in paper with description, time-period and data type.*

| ***Data Name*** | ***Paper Name*** | ***Short Name*** | ***Time Period (t_X)*** | ***Description*** | ***DAG node*** | ***Included in RFE*** |
|---|---|---|---|---|---|---|
| *AgeGroup* | Age group | - | t-1 | Infant <2, Pre-School 2 to <5, School 5 to <17 | Demographics | Yes |
| *Ethnicity* | Ethnicity | - | t-1 | Aboriginal or not (referred to as OT) | Demographics | Yes |
| *gender* | Gender | - | t-1 | Male or Female | Demographics | Yes |
| *Residence* | Residence | - | t-1 | Local or Remote | Demographics | Yes |
| *weight* | Weight | - | t-1 | Weight (kg) | Demographics | Yes |
| *enviro_exposure* | Environmental exposure | - | t-1 | Exposure to Cattle, Cats, Dogs, Goats, Sheep, Pigs, Birds, Reptiles or Other | Disease Related Risk Factors | Yes |
| *fracture_bones* | Bone fractures | - | t-1 | Current or previous fracture of involved bone(s) | Disease Related Risk Factors | Yes |
| *git_infection* | Preceeding gastro-intestinal infection | Preceeding GITI | t-1 | GITI preceeding 1 month | Disease Related Risk Factors | Yes |

| | | | | | | |
|---|---|---|---|---|---|---|
| *hx_travel* | Proceeding travel | - | t-1 | Oversease travel in proceeding 1 month | Disease Related Risk Factors | Yes |
| *hx_ssti* | Historical skin and soft tissue infection | Historical SSTI | t-1 | Historical skin and soft tissue infection (SSTI) in other family members? | Disease Related Risk Factors | Yes |
| *hx_surgery* | Preceeding surgery | - | t-1 | Surgery preceeding 1 month | Disease Related Risk Factors | Yes |
| *immunisations_utd* | Up to date immunisations | - | t-1 | Are patient's immunisations up to date | Disease Related Risk Factors | Yes |
| *immunosuppression* | Immunosuppressed | - | t-1 | Is patient immunosuppressed including receving steroids or dexamethasone | Disease Related Risk Factors | Yes |
| *inoculating_injury* | Inoculating injury | - | t-1 | Does patient have inoculating injury | Disease Related Risk Factors | Yes |
| *lrti* | Preceeding lower respiratory tract infection | Preceeding LRTI | t-1 | LRTI preceeding 1 month | Disease Related Risk Factors | Yes |
| *postop_infection* | Postoperative infection | - | t-1 | Postoperative infection including surgical site infections or deep collections relating to previous surgery | Disease Related Risk Factors | Yes |

| | | | | | | |
|---|---|---|---|---|---|---|
| *skeletal_disorder* | Skeletal disorder | - | t-1 | Does patient have a skeletal disorder | Disease Related Risk Factors | Yes |
| *tb_exposure* | Tuberculosis exposure | TB exposure | t-1 | Patient exposed to TB | Disease Related Risk Factors | Yes |
| *trauma* | Preceeding trauma | - | t-1 | Subjective preceeding trauma to the affected bone/joint in the past month | Disease Related Risk Factors | Yes |
| *urti* | Preceeding upper respiratory tract infections | Preceeding URTI | t-1 | URTI preceeding 1 month | Disease Related Risk Factors | Yes |
| *uti* | Preceeding Urinary Track Infection | Preceeding UTI | t-1 | UTI preceeding 1 month | Disease Related Risk Factors | Yes |
| *chronic_liver* | Chronic Liver disease | - | t-1 | Presence of Chronic Liver Disease | Other comorbidities | No |
| *chd* | Congenital Heart Disease | CHD | t-1 | Presence of Congenital Heart Disease | Other comorbidities | Yes |
| *diabetes* | Diabetes | - | t-1 | Presence of diabetes | Other comorbidities | Yes |
| *ear_disease* | Ear disease | - | t-1 | Chronic suppurative otitis medi or Acute otitis media | Other comorbidities | Yes |
| *head_lice* | Head lice | - | t-1 | Presence of head lice | Other comorbidities | Yes |
| *develop_delay* | Developmental delay | - | t-1 | Presence of developmental delay | Other Risk Factors | Yes |
| *malignancy* | Malignancy | - | t-1 | Patient has malignancy | Other Risk Factors | Yes |

| | | | | | | |
|---|---|---|---|---|---|---|
| *malnutrition* | Malnutrition | - | t-1 | 0-5yrs: weight-for-height at least <= -2 OR >5yrs: BMI z score <= -1 | Other Risk Factors | Yes |
| *prematurity* | Prematurity | - | t-1 | Prematurity <37 weeks | Other Risk Factors | Yes |
| *days_symptoms* | Presentation symptom duration | - | t-1 | Days of symptoms prior to presentation | Proceeding History | Yes |
| *erythema_skin* | Erythema to skin or joint at presentation | Presentation Erythema | t-1 | Erythema to skin/joint | Proceeding History | Yes |
| *joint_immobility* | Joint immobility at presentation | - | t-1 | Did patient have joint immobility at presentation | Proceeding History | Yes |
| *limb_movement* | Reduced limb movement at presentation | - | t-1 | Reduced limb movement/limb asymmetry | Proceeding History | Yes |
| *localised_swelling* | Localised swelling at presentation | - | t-1 | Presence of localised swelling at presentation | Proceeding History | Yes |
| *warmth_overlying* | Warmth overlying skin at presentation | - | t-1 | Presence of warmth overlying skin at presentation | Proceeding History | Yes |
| *BloodCultured* | Blood cultured | - | t0 | Blood culture conducted | Culture Investigation | Yes |
| *OthersCultured* | Others cultured | - | t0 | Any other culture conducted including bone, fluid or joint | Culture Investigation | Yes |

| | | | | | | |
|---|---|---|---|---|---|---|
| *WoundCultured* | Wound cultured | - | t0 | Wound swab culture conducted | Culture Investigation | Yes |
| *ul_ll* | Upper or lower limb infection | UL-LL | t0 | Infection location in upper limb or lower limb | Diagnosis of site | Yes |
| *scabies* | Scabies | - | t0 | Presence of scabies | Disease Related Comorbidities | Yes |
| *ssti* | Concurrent skin and soft tissue infection | Concurrent SSTI | t0 | Presence of SSTI at admission | Disease Related Comorbidities | Yes |
| *ssti_comorbidity* | Alternate skin and soft tissue infection | Alt SSTI | t0 | Presence of SSTI | Disease Related Comorbidities | Yes |
| *bp* | Blood Pressure | bp | t0 | Blood pressure at admission mm Hg / mm Hg | Inflammation Indicators | Yes |
| *pr* | Pulse Rate | pr | t0 | Pulse rate at admission | Inflammation Indicators | Yes |
| *temp_1* | Earliest Temp | - | t0 | Temperature at admission or peripheral centre whichever taken earlier | Inflammation Indicators | Yes |
| *temperature* | Temp at admission | - | t0 | Temperature at admission | Inflammation Indicators | Yes |
| *erythema_skin_joint* | Erythema to skin or joint at admission | Admission Erythema | t0 | Erythema to skin/joint | Other symptoms | Yes |

| | | | | | | |
|---|---|---|---|---|---|---|
| *ex_movement* | Reduced limb movement | - | t0 | Presence of reduced limb movement | Other symptoms | Yes |
| *joint_immobility1* | Joint immobility at admission | - | t0 | Presence of joint immobility at admission | Other symptoms | Yes |
| *local_swelling* | Localised swelling at admission | - | t0 | Presence of localised swelling at admission | Other symptoms | Yes |
| *rr* | Respiratory rate | - | t0 | Respiratory rate | Other symptoms | Yes |
| *septic_shock* | Septic shock | - | t0 | Presence of septic shock | Other symptoms | Yes |
| *abx_48hrs_prior* | Preceeding antibiotics | Preceeding ABX | t0 | Abx in 48 hours prior to admission | Proceeding History | Yes |
| *admitting_team* | Admitting team | - | t0 | Team that patient was admitted to with ordinal encoding | Proceeding History | Yes |
| *kingae_pcr* | Kingella Kingae PCR | Kingae PCR | t1 | PCR for Kingella. Yes, No or Not done | - | Yes |
| *BloodEvidence* | Blood evidence | - | t1 | Blood culture evidence, S. aureus, Non. S. aureus or Unculturable | Culture Results | Yes |
| *OthersEvidence* | Others evidence | - | t1 | Other culture evidence. If any S.aureus then S. aureus. If no S. aureus and Non. S. aureus then Non. S. aureus. Else Unculturable | Culture Results | Yes |

| | | | | | | |
|---|---|---|---|---|---|---|
| *WoundEvidence* | Wound evidence | - | t1 | Wound culture evidence, S. aureus, Non. S. aureus or Unculturable | Culture Results | Yes |
| *alp* | Alkaline Phosphatase at admission | Admission ALP | t1 | Alkaline Phosphatase at admission | Inflammation Indicators | Yes |
| *anc* | Absolute neutrophil count at admission | Admission ANC | t1 | Absolute neutrophil count at admission | Inflammation Indicators | Yes |
| *cr* | Creatinine at admission | - | t1 | Amount of creatinine in blood (mg/dL) | Inflammation Indicators | Yes |
| *crp_admission* | C-reactive protein at admission | Admission CRP | t1 | C-reactive protein at admission | Inflammation Indicators | Yes |
| *esr* | Erythrocyte sedimentation rate at admission | Admission ESR | t1 | Erythrocyte sedimentation rate at admission | Inflammation Indicators | Yes |
| *wcc* | White Blood Cell Count at admission | Admission WCC | t1 | White blood cell count at admission | Inflammation Indicators | Yes |
| *bony_sites_appendicular_axial* | Bone Sites | - | t2 | Locations of bone infections - list of different bones with yes or no for each | Diagnosis of site | No |
| *disease_classification* | Disease Classification | - | t2 | Osteomyelitis, Septic Arthritis, combination or | Diagnosis of site | No |

| | | | | | | |
|---|---|---|---|---|---|---|
| | | | | some other osteoarticular related infection | | |
| *number_bony_sites* | Number of Bone sites | - | t2 | Number of different bones or parts of bone infected | Diagnosis of site | No |
| *number_joints* | Number of Joint sites | - | t2 | Number of different joints or parts of joints infected | Diagnosis of site | No |
| *skeleton* | Skeleton System | - | t2 | Which part of the skeletal system is infected (Appendicular, Axial or combined) | Diagnosis of site | No |
| *duration_iv* | Duration of IV Therapy | - | t2 | Duration of IV therapy (days) | Discharge Outcomes | No |
| *icu_admin* | Admission to ICU | - | t2 | Was patient admitted to ICU | Discharge Outcomes | No |
| *length_of_stay_total* | Total Hospital Days | - | t2 | Total number of days at hospital | Discharge Outcomes | No |
| *num_surgeries* | Number of surgeries | - | t2 | Number of non-diagnostic surgical procedures | Discharge Outcomes | No |
| *resolution_discharge* | Resolution of symptoms at discharge | - | t2 | Resolution of symptoms at discharge | Discharge Outcomes | No |
| *causes_bji* | Attributable causes | - | t2 | Attributable causes of infection | Discharge Outcomes | No |

| | | | | | | |
|---|---|---|---|---|---|---|
| *duration_abx* | Duration of Proceeding Antibiotics | - | t2 | Duration of any proceeding therapy (days) | Proceeding History | No |
| *abx_reinstated* | Antibiotics Reinstated | - | t3 | Was therapy reinstated after step down | Ongoing Symptoms | No |
| *amount_school_missed* | Amount of School Missed | - | t3 | Days of school missed | Ongoing Symptoms | No |
| *documented_relapse* | Relapse | - | t3 | Was there a relapse in symptoms between discharge and the follow up period | Ongoing Symptoms | No |
| *functional_cde* | Functional Code | - | t3 | Functional code at Follow up | Ongoing Symptoms | No |
| *mobility_aid* | Mobility Aid Required +12 months | - | t3 | Was Mobility Aid Required After more than 12 months | Ongoing Symptoms | No |

### S2 Univariate analysis of progression to complicated disease by background factors

Table S2 shows the positive likelihood ratio for complications given certain background factors. The highest individual factor was having a Wound Evidence return *S. Aureus* while the lowest was being in the Infant Age Group.

*Table S2: Background factors of total study cohort and by progression to complicated disease or not*

| Background factors | Total Cohort | Background factors | Total Cohort | Background factors | Total Cohort |
|---|---|---|---|---|---|
| **Aboriginal and/or Torres Strait Islander, n (%)** | 37 (19) | 17 (25) | 20 (16) | 1.59 [0.89 2.82] | 0.89 [0.76 1.04] |
| **BloodCultured - Yes, n (%)** | 144 (75) | 50 (75) | 94 (75) | 0.99 [0.84 1.18] | 1.02 [0.61 1.71] |
| **Blood Evidence - S. aureus, n (%)** | 35 (18) | 18 (27) | 17 (14) | 1.98 [1.09 3.57] | 0.85 [0.72 0.99] |
| **Blood Evidence - Unculturable, n (%)** | 104 (54) | 31 (46) | 73 (58) | 0.79 [0.59 1.07] | 1.29 [0.95 1.75] |
| **Wound Cultured - Yes, n (%)** | 29 (15) | 21 (31) | 8 (6) | 4.9 [2.29 10.45] | 0.73 [0.62 0.87] |
| **Wound Evidence - S. aureus, n (%)** | 18 (9) | 15.0 (22) | 3.0 (2) | 9.33 [2.8 31.08] | 0.8 [0.7 0.91] |
| **Wound Evidence - Non. S. aureus, n (%)** | 10 (5) | 5.0 (7) | 5.0 (4) | 1.87 [0.56 6.22] | 0.96 [0.89 1.04] |
| **Others Cultured - Yes, n (%)** | 59 (31) | 38 (57) | 21 (17) | 3.38 [2.17 5.26] | 0.52 [0.39 0.69] |
| **Others Evidence - S. aureus, n (%)** | 42 (22) | 34.0 (51) | 8.0 (6) | 7.93 [3.9 16.14] | 0.53 [0.41 0.67] |

| | | | | | |
|---|---|---|---|---|---|
| **Others Evidence - Non. S. aureus, n (%)** | 9 (5) | 4.0 (6) | 5.0 (4) | 1.49 [0.41 5.37] | 0.98 [0.91 1.05] |
| **Prematurity, n (%)** | 20 (10) | 4 (6) | 16 (13) | 0.47 [0.16 1.34] | 1.08 [0.99 1.18] |
| **Agegroup - Infant, n (%)** | 57 (30) | 8 (12) | 49 (39) | 0.3 [0.15 0.6] | 1.45 [1.23 1.71] |
| **Agegroup - Preschool, n (%)** | 34 (18) | 11 (16) | 23 (18) | 0.89 [0.46 1.72] | 1.02 [0.9 1.17] |
| **Scabies, n (%)** | 3 (2) | 1 (1) | 2 (2) | 0.93 [0.09 10.1] | 1.0 [0.96 1.04] |
| **Upper Limb Infection, n (%)** | 36 (19) | 15 (22) | 21 (17) | 1.33 [0.74 2.41] | 0.93 [0.8 1.08] |
| **Intravenous Antibiotics Not Reinstated, n (%)** | 178 (93) | 55.0 (82) | 123.0 (98) | 0.83 [0.74 0.94] | 11.19 [2.58 48.55] |
| **URTI Preceeeding 1 Month, n (%)** | 89 (46) | 20 (30) | 69 (55) | 0.54 [0.36 0.81] | 1.57 [1.22 2.01] |
| **Immunisations Up to Date, n (%)** | 169 (88) | 58.0 (87) | 111.0 (89) | 0.97 [0.87 1.09] | 1.2 [0.55 2.62] |

### S3Recursive feature with alternate scoring function

Main text results presented a variable set that was optimised with respect to maximising the AUC score of a logistic regression model. Alternative scoring functions are possible. Using a negative log loss (higher is better) scoring function yielded similar results. Figure S1 shows the same results as in main text but with the alternate scoring function. The final ROC curve is almost identical while the elimination curve in Figure S1 is largely similar.

The resulting optimal variable set was quite similar. The main text presents the optimal set as including: : **Others evidence, up to date immunisations, bone fractures, postoperative infection, concurrent skin and soft tissue infection (SSTI), respiratory rate**, **joint immobility at admission and creatinine at admission**. The set found by the log-loss function was: **Others Evidence, Admitting Team, Immunosuppression, Up to date immunisations, Bone fractures, Malnutrition, Postoperative infection, SSTI, Joint immobility, Diastolic Blood Pressure, and Creatinine at admission**. Table S3 shows the optimal logistic regression model as in text but with the log-loss scoring function results.

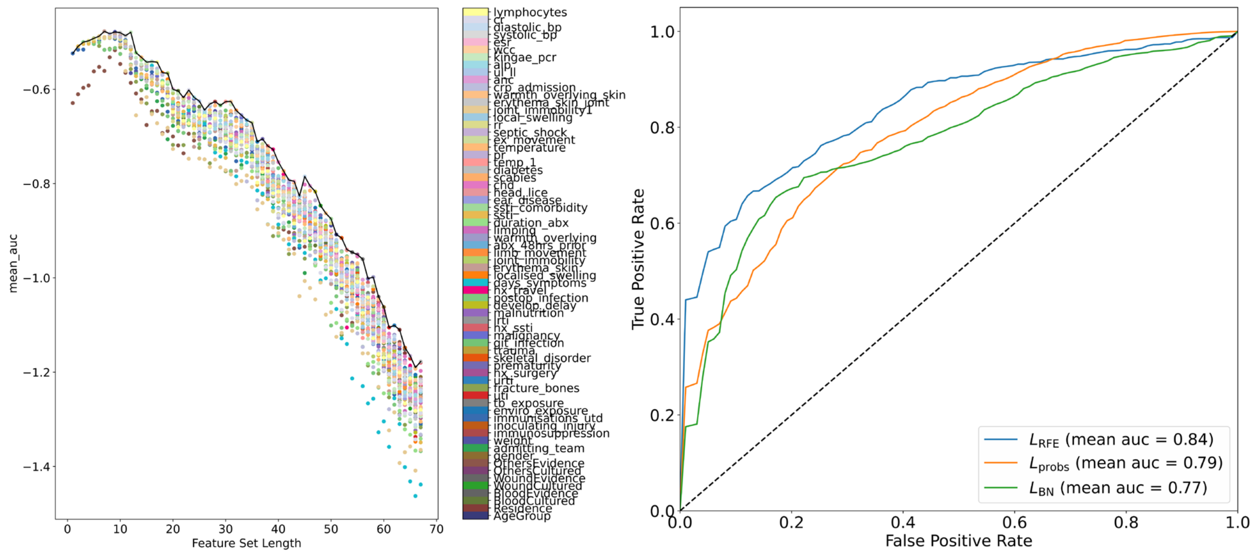


*Figure S1: Recursive feature elimination results on WARSABI dataset using log-loss instead of ROC-AUC as presented in text.* ***Left****: Mean AUC over 10 iterations of 5-folds feature reduction leads to the following score against the size of the remaining feature set. Each scatter point gives the performance of the model after a specific variable was removed, while the black line shows the best performing feature set for each feature set length. The scatter points that lie on the line reflect the variable that was chosen for removal by the recursive feature elimination algorithm.* ***Right****: ROC curves comparing the performance of the model trained on the optimal feature set, $L_{RFE}$; performance of the model trained on the BN variables, $L_{BN}$; and the performance of the model trained on the causative pathogen probabilities, $L_{probs}$.*

*Table S3: Recursive feature elimination results on WARSABI dataset using log-loss instead of ROC-AUC as presented in text.*

| ***Variable*** | $L_{RFE}$ ***OR [95% CI]*** |
|---|---|
| **Intercept** | 0.44 [0.22 1.88] |
| **Admitting Team** | |
| *admitting_team_1* | 0.72 [0.41 3.05] |
| *admitting_team_2* | 0.67 [0.38 22.93] |
| *admitting_team_3* | 1.34 [0.5 51545.8] |
| *admitting_team_4* | 1.09 [0.36 5.05] |
| *admitting_team_5* | 1.89 [0.73 10.73] |
| *admitting_team_6* | 1.29 [0.64 23960830000] |
| **Creatinine at admission** | 1.51 [0.89 2.56] |
| **Diastolic Blood Pressure** | 0.95 [0.55 1.66] |
| **Bone Fractures** | |
| *Yes* | 1.52 [0.97 2.42] |
| *No* | 0.66 [0.43 1.04] |
| **Up to date immunisations** | |
| *Yes* | 0.81 [0.57 1.16] |
| *No* | 0.96 [0.65 1.4] |
| *Not recorded* | 1.69 [0.61 4.76] |
| **Immunosuppression** | |
| *Yes* | 0.73 [0.36 74.71] |
| *No* | 1.38 [0.68 109.51] |
| **Joint Immobility** | |
| *Yes* | 1.25 [0.86 1.81] |
| *No* | 1.3 [0.87 1.95] |
| *Not recorded* | 0.56 [0.29 1.1] |
| **Malnutrition** | |
| *Yes* | 0.85 [0.41 3.31] |

| | |
|---|---|
| *No* | 1.02 [0.59 2.15] |
| *Not recorded* | 1.28 [0.62 2.97] |
| **Others *Evidence*** | |
| *Not recorded* | 0.86 [0.39 2.34] |
| *No Growth* | 0.55 [0.19 1.59] |
| *Non. Staph. Aureus* | 0.81 [0.46 1.47] |
| *Not Tested* | 0.71 [0.5 1.01] |
| *S. Aureus* | 2.21 [1.51 3.24] |
| **Historical Bone Fracture** | 0.64 [0.34 1.2] |
| *Yes* | 1.91 [0.83 4.45] |
| *No* | 0.95 [0.6 1.5] |
| **SSTI** | |
| *Yes* | 1.23 [0.86 1.77] |
| *No* | 0.71 [0.52 0.97] |
| *Not recorded* | 1.22 [0.88 1.7] |
